# AETIONOMY, a Cross-Sectional Study Aimed at validating a new taxonomy of Neurodegenerative Diseases: Study design and subject characteristics

**DOI:** 10.1101/19004804

**Authors:** Jean Christophe Corvol, Sarah Bujac, Stephanie Carvalho, Bethan Clarke, Jacqueline Marovac, Graziella Mangone, Olivier Rascol, Wassilios G Meissner, Eloi Magnin, Alexandra Foubert-Samier, Hélène Catala, Ioanna Markaki, Panagiota Tsitsi, Raquel Sanchez-Valle, Michael T. Heneka, Jose-Luis Molinuevo, Ullrich Wuellner, Per Svenningsson, Phil Scordis, Martin Hofmann-Apitius, on behalf of the AETIONOMY Clinical Consortium

## Abstract

**Background:** Although advances in the understanding of neurodegenerative diseases (NDDs) have led to improvements in classification and diagnosis and most importantly to new therapies, the unmet medical needs remain significant due to high treatment failure rates. The AETIONOMY project funded by the Innovative Medicine Initiative (IMI) aims at using multi-OMICs and bioinformatics to identify new classifications for NDDs based on common molecular pathophysiological mechanisms in view of improving the availability of personalised treatments.

**Objectives:** The purpose of the AETIONOMY cross-sectional study is to validate novel patient classification criteria provided by these tools.

**Methods:** This was a European multi centre, cross-sectional, clinical study conducted at 6 sites in 3 countries. Standardised clinical data, biosamples from peripheral blood, cerebrospinal fluid, skin biopsies, and data from a multi-OMICs approach were collected in patients suffering from Alzheimer’s and Parkinson’s disease, as well as healthy controls.

**Results:** From September 2015 to December 2017 a total of 421 participants were recruited including 95 Healthy Controls. Nearly 1,500 biological samples were collected. The study achieved its objective with respect to Parkinson’s disease (PD) recruitment, however it was unable to recruit many new Alzheimer Disease (AD) patients. Overall, data from 413 evaluable subjects (405 PD and 8 AD) are available for analysis. PD patients and controls were well matched with respect to age (mean 63.4 years), however, close gender matching was not achieved. Approximately half of all PD patients and one At-Risk subject were taking dopamine agonists; rates of Levodopa usage were slightly higher (∼60%). Median MDS-UPDRS Part III Scores (OFF state) ranged from 45 (SD 18) in those with Genetic PD to 2 (SD 3) in Healthy Controls. The standardised methodologies applied resulted in a high-quality database with very few missing data.

**Conclusion:** This is one of the collaborative multi-OMICs studies in individuals suffering from PD and AD involving a control group. It is expected that the integration of data will provide new biomarker-led descriptions of clusters of patient subgroups.

## Introduction

Dementia, mainly represented by Alzheimer’s disease (AD), affects 44 million people globally, and that figure is set to rise to 135 million by 2050, mostly due to the ageing of the population.^1^ Meanwhile an estimated 4-6 million people globally suffer from Parkinson’s disease (PD), the second cause of neurodegenerative disease.^2^ There is no cure for these devastating diseases, and caring for patients as their disease progresses represents an immense burden for family members, carers, and health and social care systems. The way these diseases are classified is hampering efforts to develop effective, targeted treatments. Currently, diseases are defined largely on the basis of the patient’s symptoms and where they occur in the body. There is growing evidence that while two patients may be classified as having the same disease, the genetic or molecular causes of their symptoms may be very different. In other cases, diseases that are currently defined as separate conditions may share a common molecular basis. There is therefore now broad recognition that the way diseases are classified needs to change, and the field of neurodegenerative diseases in particular is considered to be ripe for a rethink.^3^

AETIONOMY was a European project funded by the Innovative Medicine Initiative which intended to develop tools to better classify patients into subgroups based on their underlying pathogenic mechanisms (http://www.aetionomy.eu). During the lifetime of the project the team explored the classification of neurodegenerative disease by dissecting molecular causes of disease and exploring links to clinical evidence. An important activity for the project was the evaluation, and ultimate organisation, of historical data from a range of sources pertaining to the field. This knowledgebase formed the genesis of mechanistic hypotheses that, alongside canonical theories for the drivers of neurodegeneration, were evaluated utilising computational^4^ and conventional laboratory methodologies. To facilitate these objectives, a prospective cross-sectional study was initiated that aimed to support the clinical validation of the identified mechanisms in both AD and PD patients. The present publication provides details of the study design, the patient population and their baseline clinical characteristics. It also highlights the study management strategy and the standardised methodologies implemented to ensure quality of the data and sample collection.

## Material and methods

### Study design

This is a European multi-centre, cross-sectional clinical study, aimed at collecting clinical, brain imaging and biological data on AD and PD patients, individuals at risk of AD and PD, and healthy controls. The primary objective of the study was to validate the new taxonomy of neurodegenerative diseases proposed by the AETIONOMY Consortium in a real cohort of patients, representative of the continuum of AD and PD. Secondary objectives were to describe the correlations between clinical features and biomarkers across neurodegenerative diseases.

### Participants

Inclusion and exclusion criteria for the two groups of patients (AD and PD) and their matching controls are described in detail in **Table S1**. For the PD group, subjects were: patients with a diagnosis of idiopathic PD according to UK PD Society Brain Brank,^5^ and a disease duration less than 10 years at inclusion; patients with autosomal dominant (*SNCA, LRRK2* or *GBA*) or autosomal recessive (*PARK2*) forms of PD; a group of subjects “at risk of PD” defined as subjects without symptom of PD but either being a first degree relatives of patients with an autosomal dominant form of PD, or subjects with polysomnography-confirmed idiopathic REM sleep behavioural disorders; healthy subjects matched for age and gender without symptoms of PD or other neurological disorders were recruited as controls. For the AD group, we initially aimed to recruit subjects with biologically confirmed AD at the clinical stage, at the prodromal stage, and matched healthy controls. However, we decided to stop enrolment of the AD group shortly after the recruitment started because another IMI-funded project was planning to build a prodromal AD cohort (European Prevention of Alzheimer’s Dementia (EPAD) Longitudinal Cohort Study (LCS), http://ep-ad.org/). The AETIONOMY study thus finally mainly focused on the PD group.

### Study procedures

Subjects were recruited in six University Hospitals in France (Pitié-Salpêtrière (Paris), Toulouse, Bordeaux, and Besançon), Germany (Bonn), and Sweden (Karolinska University Hospital, Stockholm). Participants were invited to attend a single study visit at the hospital, during which standardised clinical assessments and biological sampling were conducted.

After obtaining informed consent and the confirmation of inclusion/exclusion criteria, clinical data were collected by a neurologist including: demographics (e.g. age, sex, ethnicity), vital exams (e.g. blood pressure, electrocardiogram), personal and familial medical history, and environmental factor exposure history. Additional disease-specific clinical and neuropsychological assessments were also performed. For the PD group (including controls) these were: Movement Disorder Society-Unified PD Rating Scale (MDS-UPDRS), Hoehn & Yahr stage, Schwab & England Activities of Daily Living (SE-ADL) scale, Non-Motor Symptoms Scale (NMSS), Hospital Anxiety and Depression Scale (HAD), 39-item Parkinson’s Disease Questionnaire (PDQ-39), REM Sleep Behaviour Disorder Questionnaire – Hong Kong (HK-RBD), Epworth Sleepiness Scale (ESS), University of Pennsylvania Smell Identification Test (UPSIT) and Stand-walk-sit test. For the AD group (including controls) these were: The 30-item Geriatric Depression Scale (GDS), State-Trait Anxiety Inventory (STAI), Pittsburgh Sleep Quality Index (PSQI), Brain MRI, FCSRT-IR (Buschke) and Clinical Dementia Rating (CDR). The Montreal Cognitive Assessment (MOCA), Mini Mental State Examination (MMSE), Frontal Assessment Battery (FAB), Repeatable Battery for the Assessment of Neuropsychological Status (RBANS) and environment questionnaire including IPAQ were administered to subjects from both groups. Neuropsychological assessments were performed during the visit by trained neuropsychologists. Clinical data were collected in an electronic case report form (eCRF) using the REDCap™ system.^6^ The eCRF was developed in collaboration with the Bioinformatics Core Facility of the Centre for Systems Biomedicine (LCSB, University of Luxembourg, UL). All data and analyses produced are stored in a project TranSMART database used and hosted by LCSB, UL and managed by the ICM.

### Biological samples

Blood samples were collected in the morning, in a fasted state, into: Serum Separating Tubes (SST, serum collection) for serum, Lithium Heparin tubes (LH) for plasma, and EDTA tubes for DNA extraction. Serum and plasma were frozen and stored at −80°C for biomarker analysis (proteomics, metabolomics, lipidomics, genomics, epigenomics). EDTA plasma samples were also snap-frozen in liquid nitrogen (N2) and stored in N2 for the analysis of neuroinflammation biomarkers. Cerebrospinal Fluid was collected by lumbar puncture. Samples were centrifuged at 4°C within 1 hour, for 10 min at 2,000g and then progressively frozen and stored at −80°C also for biomarker analysis (proteomics, metabolomics, lipidomics). A subset of samples was snap-frozen immediately (within 60 seconds) and stored in liquid N2 for the analysis of neuroinflammation biomarkers. Skin biopsies were also proposed for fibroblast preparations frozen and stored in 10% dimethyl sulfoxide (DMSO) medium. Lumbar puncture and skin biopsy were optional for the participants.

Samples were collected, processed and stored locally in each site following the established standard operating procedures. Samples were then distributed in different centres of the Consortium for specific analyses: genomic analysis (ICM), epigenetics (UKB), proteomics (KI), neuroinflammatory markers (UKB), lipidomics (UKB), AD markers (IDIBAPS), and markers of autophagy and insulin resistance (SARD). Sample management procedures were established prior to the start of the project and clinical centres were trained to ensure standardised collection, preparation and transportation of the samples across countries. All the remaining samples were finally centralised at the ICM biobank (Banque ADN et Cellules – ICM).

### Regulatory and ethical approvals

The sponsor of the study was the French institution *Institut National de la Santé et de la Recherche Médicale* (INSERM). We took advantage of the European Clinical Research Infrastructure Network (ECRIN) which took care of all regulatory approvals and applications, under the responsibility of the sponsor, in other European countries, i.e. in Germany and Sweden. The study protocol was approved by the Ethical Review Boards (ERB) of the three participating countries. The study was conducted in accordance with the standards set by International Conference on Harmonization and Good Clinical Practice (ICH-GCP), and to the ethical principles that have their origin in the Declaration of Helsinki (2013). The protection of the confidentiality of records that could identify the included subjects is ensured as defined by the EU Directive 2001/20/EC and the applicable national and international requirements relating to data protection in each participating country.

### Sample size calculation

The study was originally designed to recruit approximately 655 subjects in two disease groups (PD and AD). This initial sample size was chosen to allow the identification of subgroups with adequate precision, in accordance with our primary criteria. The primary objective aimed at identifying subgroups of subjects that correspond to a hypothetic causative biological pathway and corresponding to a particular biomarker. Depending on the hypothesis to be validated, the size of the corresponding subgroup may be highly variable. For example, a subgroup defined by genetics could represent only 2% of the total sample. By contrast, a subgroup defined by non-omics markers may consist of 30% to 40% of the total population. The sample size was calculated based upon the idea that we identify a subgroup in X% of the population and we want to measure X% with a pre-defined precision (supplementary material, **Table S2**). Sample-size was estimated using standard software (nQuery. Sample Size and Power Calculation. “Statsols” (Statistical Solutions Ltd), Cork, Ireland) based upon the length of a 95% confidence interval (CI) around the size of the sub-group being “shorter” or “longer”. The shorter confidence interval means a more precise estimate and requires more patients. For example, a subgroup consisting of 2.5% of the sample, such as may be found for a subgroup defined by genetics, will have a “reasonable” 95% confidence limit of (1.3%, 3.7%) when 655 subjects are studied.

### Statistical analysis

Key demographic, clinical, and disease severity variables are summarised in a descriptive manner only. The intention is simply to provide an overview of the key information that have been collected for each subgroup and to point out some of the main features of this data. Statistical modelling is beyond the scope of this particular manuscript and formal statistical comparisons of the groups are not included. Summary statistics of subject data at entry were produced for each PD subgroup using PC SAS 9.4 (SAS Institute Inc., Cary, NC, USA) and the statistical tools within tranSMART. Due to the low subject numbers, summaries are not provided for the AD patients, nor their controls. Continuous variables are described using means and standard deviations or medians with interquartile ranges where non-gaussian distributions were suspected. Numbers and percentages are provided for each categorical variable. Unless specified otherwise, percentage denominators are the number of non-missing values.

## Results

### Enrolment of participants

A total of 421 subjects were screened for the study. Eight of these subjects were excluded from the final dataset. The reasons for exclusion were: screen failure (n=3), participant withdrawal upon investigator decision (n=2) and subject decision (n=3) (supplementary material, **Figure S1**). One serious Adverse Event was reported that related to a study procedure (a post-lumbar puncture syndrome with a complete recovery). All data from the subject who experienced this SAE was included in the final analysis set. Overall, data from 413 subjects were available for analysis: 405 in the PD group and 8 in the AD group (**Table 1**). The study achieved its overall objective with respect to the PD group recruitment (412 screened vs. 415 planned, including controls), although the numbers of patients recruited with Genetic PD subjects was fewer than desired (25 screened vs 40 planned). The “at risk PD” subgroup included 25 subjects with idiopathic RBD and 14 with a first degree relative of a genetic PD index case. Due to the early termination of AD recruitment, the number of AD subjects is very small (n=8) and data from this group are not summarised in any further detail in this manuscript. In total, nearly 1500 biological samples were collected (**Table 2**). Biomarker analyses of these samples is currently being undertaken and will be presented in subsequent AETIONOMY Consortium manuscripts.

**Table 1:** Study populations.

| AETIONOMY<br>Cross-sectional study<br>subgroups | Idiopath<br>ic PD<br>patients | PD group |  |  |  |  |  |  | AD group |  |  |  |
| --- | --- | --- | --- | --- | --- | --- | --- | --- | --- | --- | --- | --- |
|  |  | Genetic PD patients |  |  |  | At Risk<br>of PD<br>subjects | Healthy<br>controls | Tota<br>l | Prodrom<br>al AD<br>subjects | At Risk<br>of AD<br>subjects | Healthy<br>controls | Tota<br>l |
|  |  | PARKIN<br>mutation | GBA<br>mutation | LRRK2<br>mutatio<br>n | SNCA<br>mutation |  |  |  |  |  |  |  |
| Subjects, screened n (%) | 255<br>(54.6) | 8 (1.9) | 6 (1.5) | 10 (2.4) | 1 (0.2) | 39 (9.5) | 93 (22.6) | 412 | 2 (22.2) | 5 (55.6) | 2 (22.2) | 9 |
| Subjects, analysed n (%) | 251<br>(62.0) | 8 (2.0) | 6 (1.5) | 10 (2.5) | 1 (0.2) | 39 (9.6) | 90 (22.2) | 405 | 2 (25.0) | 4 (50.0) | 2 (25.0) | 8 |

The Parkinson's disease (PD) patient population of AETIONOMY Cross-Sectional Study was divided into three subgroups: Idiopathic PD patients, Genetic PD patients and patients At Risk of PD. The Alzheimer's disease (AD) patient population was composed of two subgroups: Prodromal AD subjects and patients At Risk of AD.
Both of the PD and AD groups of patients were matched with healthy control populations.

**Table 2:**
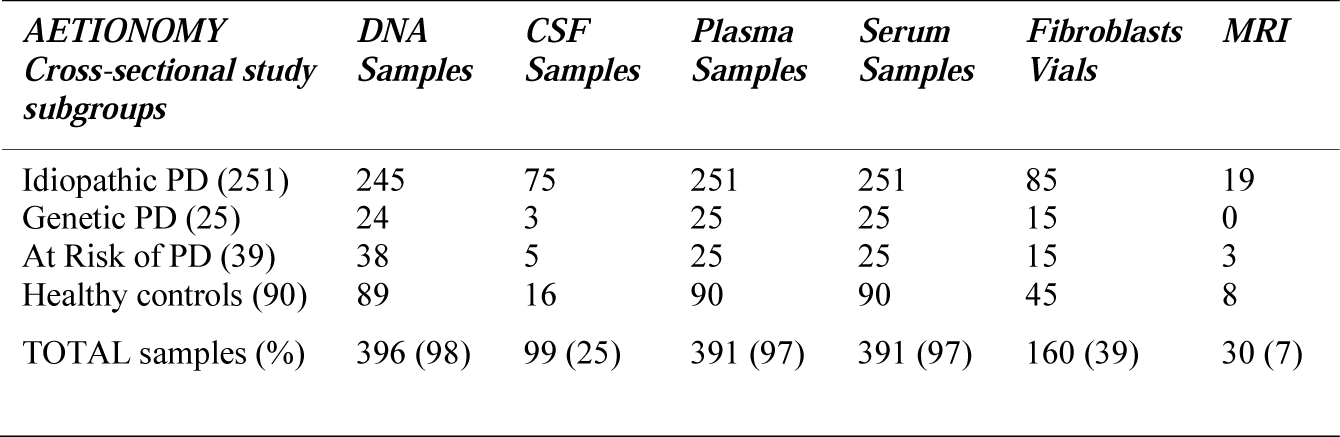
The AETIONOMY-CS PD biological collection.

### Demographic and environmental characteristics

Demographic and other subject characteristics are summarised in **Table 3**. Data ascertainment was extremely high for all the variables, as can be seen from the percentages in the final column. PD patients and controls were well matched with respect to age, however close gender matching was not achieved, with the Healthy Control group containing a much higher proportion of females (66%) compared to both the PD and the At-Risk groups (maximum 44% female). The majority of subjects in each group were Caucasian/White (range: 76% in Genetic PD to 98% Idiopathic PD).

**Table 3:**
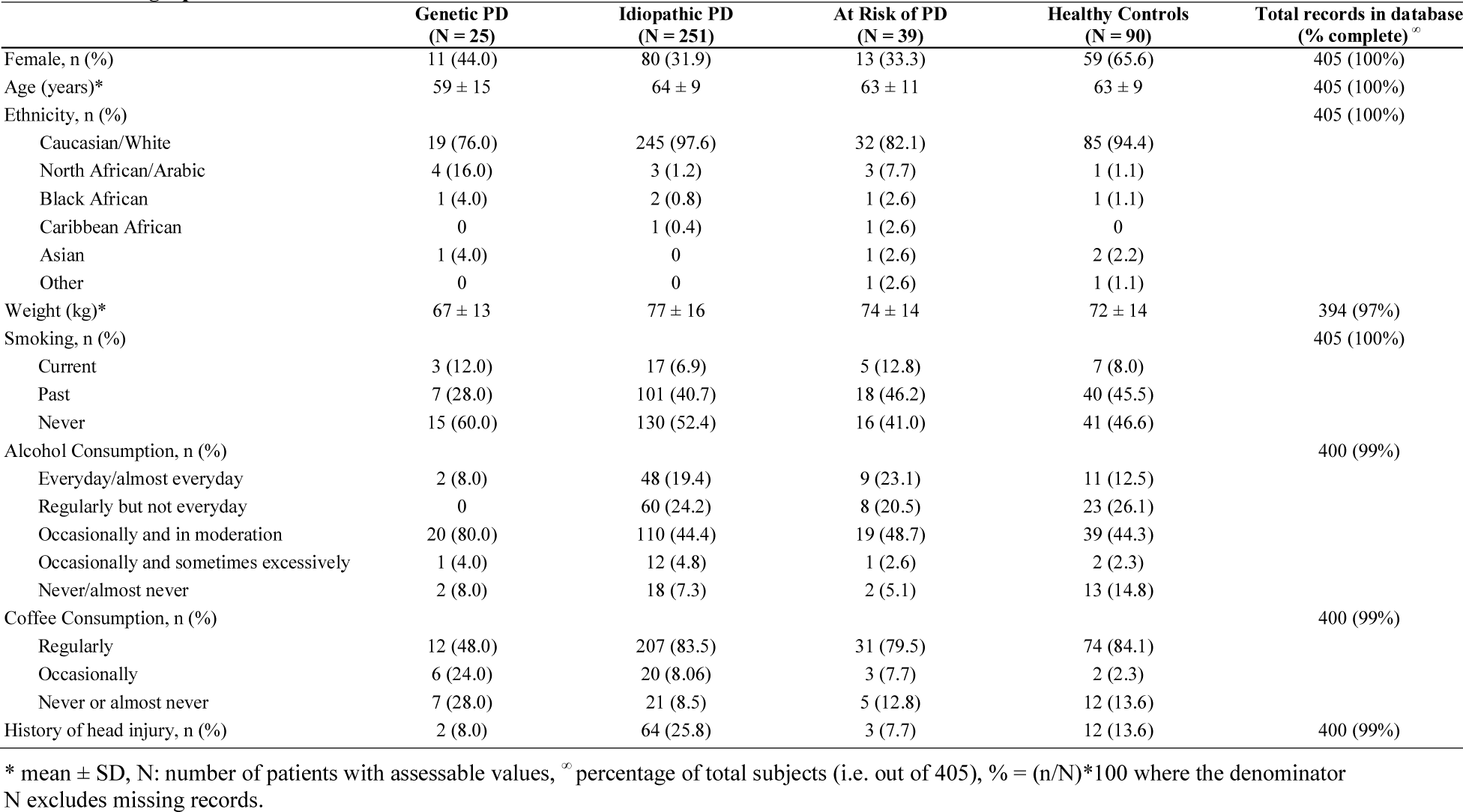
Demographic and other characteristics.

Amongst the key environmental factors, rates of current smoking (a protective factor for PD) were low (<13%) across all groups, with more than half of the PD patients having never smoked. The proportion of those who consumed coffee was observed to be much lower in the Genetic PD Group (48%) compared to all other groups where the rates were over 80%. As expected, a history of head injury was most common in the Idiopathic PD group.

### Clinical characteristics

Clinical characteristics, disease severity scores and medication use are summarised in **Table 4**. As expected, the majority (64%) of subject with Genetic PD and very few Idiopathic PD subjects or controls (6% for each) reported a family history of PD. Over a third (40%) of the At-Risk PD group also reported a family history of PD. Subjects with Genetic PD had a lower average age at onset (45 compared to 61 years) and a much longer average disease duration (144 compared to 28.5 months) compared with Idiopathic PD subjects.

**Table 4:** Clinical and disease characteristics.

|  | **Genetic PD**<br>**(N = 25)** | **Idiopathic PD**<br>**(N = 251)** | **At Risk of PD**<br>**(N = 39)** | **Healthy Controls**<br>**(N = 90)** | **Total records in database**<br>**(% complete)<sup>∞</sup>** |
| --- | --- | --- | --- | --- | --- |
| Family history of PD (1<sup>st</sup>/2<sup>nd</sup> degree), n (%) | 16 (64.0) | 14 (5.6) | 15 (39.5) | 5 (5.8) | 398 (98%) |
| Age at onset (years)* | 45 ± 16 | 61 ± 9 | N/A | N/A | 274 (99%)<sup>§</sup> |
| Disease duration since diagnosis (months)<sup>+</sup> | 144 (125) | 28.5 (43) | N/A | N/A | 270 (98%)<sup>§</sup> |
| Concomitant Medication, n (%) | 23 (92%) | 237 (94%) | 30 (77%) | 48 (55%) | 403 (99%) |
| PD Medication, n (%) | 20 (80%) | 198 (79%) | 1 (3%) | 0 | 405 (100%) |
| Dopamine agonist, n (%) | 13 (52) | 127 (51) | 1 (3) | 0 | 405 (100%) |
| Dopamine agonist LEDD (mg)<sup>+</sup> | 160 (140) | 157 (142) | 98 (0) | N/A |  |
| Levodopa, n (%) | 15 (60) | 143 (57) | 0 | 0 | 405 (100%) |
| Levodopa LEDD (mg)<sup>+</sup> | 532 (825) | 350 (232) | N/A | N/A |  |
| Total LEDD (mg)<sup>+</sup> | 944 (429) | 496 (394) | 98 (0) | 0 |  |
| MDS-UPDRS Score* |  |  |  |  |  |
| Part I | 13 ± 7 | 9 ± 5 | 7 ± 3 | 3 ± 3 | 397 (98%) |
| Part II | 15 ± 9 | 9 ± 5 | 1 ± 2 | 1 ± 1 | 397 (98%) |
| Part III (OFF State) | 45 ± 18 | 30 ± 15 | 10 ± 5 | 2 ± 3 | 321 (79%) |
| Part IV | 6 ± 4 | 1 ± 3 | 0 ± 0 | 0 ± 0 | 391 (97%) |
| Schwab and England Score, n (%) |  |  |  |  | 391 (97%) |
| ≤ 50% | 3 (12.5) | 2 (0.8) | 0 | 0 |  |
| 60% or 70% | 3 (12.5) | 10 (4.0) | 0 | 0 |  |
| 80% or 90% | 17 (70.8) | 202 (81.1) | 6 (16.2) | 3 (3.4) |  |
| 100% | 1 (4.2) | 35 (14.1) | 31 (83.8) | 85 (96.6) |  |
| Hoehn and Yahr Score, n (%) |  |  |  |  | 391 (97%) |
| 0 | 0 | 5 (2.1) | 33 (84.6) | 83 (98.8) |  |
| 1 | 0 | 34 (13.9) | 0 | 0 |  |
| 2 | 17 (70.8) | 156 (63.9) | 6 (15.4) | 1 (1.2) |  |
| 3 | 2 (8.3) | 46 (18.9) | 0 | 0 |  |
| 4 | 0 | 3 (1.2) | 0 | 0 |  |
| 5 | 5 (20.8) | 0 | 0 | 0 |  |
| Motor Fluctuations, n (%) | 18 (72.0) | 35 (14.1) | 0 | 0 | 391 (97%) |
| Dyskinesia, n (%) | 18 (72.0) | 23 (9.3) | 0 | 0 | 391 (97%) |
| HADS |  |  |  |  | 395 (98%) |
| Anxiety Score* | 8 ± 4 | 6 ± 4 | 7 ± 3 | 5 ± 3 |  |
| Depression Score* | 5 ± 3 | 4 ± 3 | 3 ± 3 | 2 ± 3 |  |
| NMSS total score* | 10 ± 5 | 9 ± 5 | 8 ± 4 | 3 ± 3 | 388 (96%) |
| MMSE total score * | 28 ± 2 | 28 ± 2 | 29 ± 1 | 29 ± 1 | 389 (96%) |
| MoCA total score* | 26 ± 4 | 26 ± 3 | 27 ± 3 | 27 ± 3 | 392 (97%) |
| RBANS total score* | 93 ± 20 | 92 ± 17 | 102 ± 14 | 101 ± 16 | 377 (93%) |

\* mean ± SD, <sup>+</sup> median (IQR), <sup>§</sup> % for PD subjects only, <sup>∞</sup> percentage of total subjects (i.e. out of 405), % = (n/N)\*100 where the denominator N excludes missing records, \*100, LEDD: Levodopa Equivalent Daily Dose, HADS: Hospital Anxiety and Depression Scale, MDS-UPDRS: Movement Disorder Society-Sponsored Revision of the Unified Parkinson's Disease Rating Scale, MMSE: Mini-Mental State Examination, MoCA: Montreal Cognitive Assessment, NMSS: Non-motor Symptoms Scale, RBANS: Repeatable Battery for the Assessment of Neuropsychological Status.

Dopamine agonists were taken by approximately half of all PD patients and one At-Risk subject (for restless leg syndrome); rates of Levodopa usage were slightly higher (∼60%). Consistent with a longer disease duration, the total average Levodopa Equivalent Daily Doses was higher in Genetic PD as compared to Idiopathic PD; the difference being accounted for by higher daily dosages of Levodopa in the Genetic PD group.

A consistent pattern was observed across a number of disease severity scales, such as MDS-UPDRS III, SE-ADL, Hoehn and Yahr, HADS Depression Score and NMSS. Genetic PD subjects tended to have the most severe scores, probably reflecting the longer average duration of their disease, compared to Idiopathic PD subjects. The data from these scales also highlighted that, whilst not as severely affected as the PD subjects, those At Risk of PD were generally more severe than Healthy Controls. As an example, average MDS-UPDRS part III scores were 45 (SD 18) in Genetic PD, 30 (SD 15) in Idiopathic PD, 10 (SD 5) in At Risk subjects and 2 (SD 3) in Healthy Controls. Importantly, MDS-UPDRS part III was performed in OFF-state to better represent the severity of the disease in treated patients. Finally, the majority of the Genetic PD (72%) group, but only a few of the Idiopathic PD group (9-14%), experienced Dyskinesia and Motor Fluctuations.

Average cognitive scores (MMSE, MOCA and RBANS) were very similar between groups. Specifically, there was minimal difference between the At-Risk group and Healthy Controls.

## Discussion

The potential for the utility of the large amount of data from multiple disparate sources (literature, public, and private databases) to pave the way for a better classification of patients, based on underlying causes instead of symptoms, was the core objective of the AETIONOMY project. Achieving this goal goes far beyond the scope of any single company or university; the key to success can only come from the broad nature of the project consortium, which brings together expertise in neurodegenerative diseases, molecular biology, clinical research, research ethics, data modelling and simulation, data standards, and patient involvement in research.

The AETIONOMY team’s focus on systematically capturing and representing knowledge on neurodegenerative diseases in a computable form to and the generation of a large inventory of multiscale (ranging from the molecular level to the clinical level and cognitive readouts) mechanistic hypotheses for AD and PD lays the foundations for clinical verification of these models in an appropriate and clinically relevant cohort of patients. The role of the AETIONOMY cross-sectional study in this context is therefore to support the validation of novel patient classification criteria provided by these approaches.

We present here the design and the main clinical characteristics of a cross-sectional cohort of subject which aim to represent the continuum of PD pathology. We believe that this clinical dataset and its associated biological collection are suitable for an attempt to stratify the disease according to its underlying mechanisms. As compared to cohorts recruiting only PD patients, our population includes idiopathic and genetic forms of PD, early and late stage of the disease, subject at the prodromal stage (idiopathic RBD) or at the preclinical stage (first degree relatives of genetic PD) of the disease, and healthy controls. Only a few initiatives, like the Parkinson’s Progression Marker Initiative (PPMI),^7^ have collected in a standardised manner clinical data and biological samples from all these subpopulations representing the whole spectrum of the disease.

In our study, clinical assessments were carefully chosen to allow a comprehensive description of the motor and the non-motor features of the disease. Demographic characteristics are very similar to what has been described in previous studies in PD or prodromal PD,^7-11^ although the age at onset is younger than in the general population, probably because patients were recruited in expert centres. As expected, at-risk individuals presented average scores in-between PD patients and healthy controls for most of the rating scales.^12^ An extensive biological collection is associated with the AETIONOMY clinical dataset including DNA, plasma, and serum for virtually all patients, CSF for 25% and fibroblasts derived from skin biopsies for 39% of the subjects.

State of the art omics experiments and/or specific biomarker analyses are currently ongoing by members of our Consortium in order to achieve the ambitious goal of stratifying PD patients based on their underlying mechanisms. It is particularly difficult for neurodegenerative diseases since no clear biomarkers have been found to be specifically related to these mechanisms. Current and past exposure to environmental factors and concomitant medication have been collected to adjust for potential confounding factors in upcoming biomarker analyses.

Our study has some important limitations that must be highlighted. Our cohort was initially designed to recruit patients across neurodegenerative diseases, including PD and AD patients. We did not reach our goal in terms of recruitment of AD patients because a competitive project IMI funded, the European Prevention of Alzheimer’s Dementia (EPAD) study, was initiated soon after AETIONOMY started. The EPAD cohort will provide complementary data to our cohort which mainly focuses on PD. Although subjects enrolled in our study represent the continuum of PD from healthy controls to subject at risk, and PD patients, AETIONOMY is a cross-sectional study. Future longitudinal prospective studies are needed to investigate the progression of the disease, and determine the different trajectories according to the underlying mechanisms. Finally, the sample size of our dataset remains relatively small to perform unbiased analyses that require very large numbers of patients to correct for multiple comparison testing. Findings from our analyses will thus require replication in independent cohorts.

In conclusion, we provide here a new cohort for investigating the complex biology of neurodegenerative diseases, focusing on PD. Beyond the specific objective of AETIONOMY, we believe that this dataset is valuable for other research on biomarkers, patient stratification, and PD mechanisms. Following the concept of open sciences, our Consortium has implemented a process for data sharing, and the data as well as the samples will be made available on request to the scientific community.

## Data Availability

All data are available on request to the corresponding author

## Funding

The research leading to these results has received support from the Innovative Medicines Initiative Joint Undertaking under grant agreement no. 115568, resources of which are composed of financial contribution from the European Union’s Seventh Framework Programme (FP7/2007-2013) and EFPIA companies’ in-kind contribution.

### Acknowledgements

This paper is presented on behalf of the AETIONOMY Clinical Consortium. It was developed with input from the AETIONOMY Steering Committee. We thank all the members of each recruiting centre for their dedicated effort, enthusiasm, promptness and care in the recruitment and assessment of the participants in this study.

The members of the AETIONOMY Clinical Consortium include (in alphabetical order of their country affiliations):

Sarah Bujac, UCB Pharma SA, Belgium

Bethan Clarke, UCB Pharma SA, Belgium

Jacqueline Marovac, UCB Pharma SA, Belgium

Phil Scordis, UCB Pharma SA, Belgium

Stephanie Carvalho, Institut du Cerveau et de la Moelle épinière, Paris, France

Jean-Christophe Corvol, Institut du Cerveau et de la Moelle épinière, Paris, France

Bruno Dubois, Institut du Cerveau et de la Moelle épinière, France

Cecile Gaudebout, Institut du Cerveau et de la Moelle épinière, Paris, France

Graziella Mangone, Institut du Cerveau et de la Moelle épinière, Paris, France

Sylvie Forlani, Banque ADN & cellules, Institut du Cerveau et de la Moelle épinière, Paris, France

Ludmila Jornea, Banque ADN & cellules, Institut du Cerveau et de la Moelle épinière, Paris, France

Philippe Martin-Hardy, Banque ADN & cellules, Institut du Cerveau et de la Moelle épinière, Paris, France

Yassaman Ghassab, Banque ADN & cellules, Institut du Cerveau et de la Moelle épinière, Paris, France

Eloi Magnin, CHU Besançon, France

Alexandra Foubert-Samier, IMNc, Hôpital Pellegrin, CHU Bordeaux, France

Brice Laurens, IMNc, Hôpital Pellegrin, CHU Bordeaux, France

Wassilios Meissner, Hopital Pellegrin Bordeaux, France

Umberto Spampinato, Hôpital Pellegrin, CHU Bordeaux, France

Sylvain Vergnet, IMNc, Hôpital Pellegrin, CHU Bordeaux, France

Olivier Rascol, CHU Toulouse, France

Claire Thalamas, CHU Toulouse, France

Monique Galitzky, CHU Toulouse, France

Fabienne Calvas, CHU Toulouse, France

Fabienne Ory-Magne, CHU Toulouse, France

Christine Brefel-Courbon, CHU Toulouse, France

Michael Heneka, Universitaetsklinikum Bonn, Germany

Pawel Tacik, Universitaetsklinikum Bonn, Germany

Ullrich Wüllner, Universitaetsklinikum Bonn, Germany

Martin Hofmann-Apitius, Fraunhofer Institute for Algorithms and Scientific Computing SCAI, Germany

José Luis Molinuevo, Barcelonabeta Brain Research Center, Spain

Mircea Balasa, Consorci Institut d’Investigacions Biomediques August Pi i Sunyer, Spain

Beatriz Bosch, Consorci Institut d’Investigacions Biomediques August Pi i Sunyer, Spain

Spain Raquel Sánchez-Valle, Consorci Institut d’Investigacions Biomediques August Pi i Sunyer, Spain

Ioanna Markaki, Karolinska Institutet, Stockholm, Sweden

Per Svenningsson, Karolinska Institutet, Stockholm, Sweden

Panagiota Tsitsi, Karolinska Institutet, Stockholm, Sweden

